# HPV and p16 expression association with 5-year survival in oral squamous cell carcinoma patients of North-East India

**DOI:** 10.1101/2023.01.17.23284649

**Authors:** Rajjyoti Das, Rupesh Kumar, Avdhesh Kumar Rai, Anupam Sarma, Lopamudra Kakoti, Amal Chandra Kataki, Mouchumee Bhattacharyya, Manoj Kalita

## Abstract

**Background:** In our study, we examined the 5-year survival of OSCC patients with HPV positive or negative status along with p16 protein expression.

**Method:** A total of 72 biopsy tissue specimens from histologically confirmed oral squamous cell carcinoma (OSCC) patients were collected. HPV detection and genotyping were performed using HPV E6/E7 and HPV-type specific multiplex primer for nested-PCR. Immunohistochemistry evaluation of pl6 was conducted. SPSS statistical software (ver 20) was used for data analysis.

**Results:** High risk-HPV (hr-HPV) DNA positivity was found in 27.7% (n=20) of OSCC patients. Stage III OSCC patients were 7.80 times more likely to survive 5 years than stage IV patients (OR-7.80 CI-95%; P-0.03). Among the hr-HPV positive OSCC patients, we found that the median survival time for the 1st year (95%), 3 years (78.5%), and 5 years (38.5%) was significantly higher than that of the hr-HPV negative [1st year (78.6%), 3 years (45.2%) and 5 years (38.5%)] OSCC patients (P-0.03 The survival of male patients with hr-HPV positive OSCC is 9.75 times greater than the survival of patients with HPV negative OSCC (OR-9.75; CI-95%; P-0.05). The p16 expression level (low to overexpression) group and negative P16 expression group of OSCC patients have not demonstrated a significant association with 5-year survival.

**Conclusion:** We conclude that in OSCC cases of North-East India, the presence of hr-HPV in OSCC cases could be a good predictor of 5-year survival rate. Expression of p16 does not appear to have any significant association with 5-year survival.

## INTRODUCTION

Globally, there are estimated to be 377,713 new cases of oral cavity and lip cancers, and 177,757 deaths, ranking it seventeenth in terms of incidence and mortality [1]. Human papillomaviruses (HPVs) are implicated in approximately 5% of all cancers [2]. There have been reports of HPV prevalence in India’s oral cavity cancers of 48%, 33.6%, 15%, 27.5%, 28% in south, eastern, western, central, and north-eastern India respectively [3-7]. Human papillomavirus (HPV) infections are considered a significant risk factor for the onset of oral cancer in young individuals in North America, Europe, and South-Central Asia [8-10]. According to national cancer registry program report, oral cancer mortality and incidence are higher in the North-East region of India than in other parts of the country [11]. A significant correlation has been shown between HPV infection and Oral Squamous Cell Carcinoma (OSCC) in the last two decades [12]. Cancer of the oral cavity has a poor prognosis at advanced stages, and metastatic lymph nodes have been found to be associated with worse survival rates even in early stages [13]. It is reported that OSCCs in stages I and II were associated with better survival rates [14].

Two hr-HPV genotypes, HPV-16 and HPV-18 are major identifiable types in OSCC [12-14]. However, the HPV-16 type constituted 90% and HPV-18 30% of HPV-associated OSCCs [15]. These hr-HPV types, 16 and 18, produce carcinogenic activity by expressing E6 and E7 oncoproteins [16]. Oncogenic activation of the E6 oncogene leads to the degradation of p53 through the E3 ubiquitin ligase E6AP, while viral E7 oncoprotein inhibit tumor-suppressor proteins p53 and retinoblastoma protein (Rb) [17]. HPV E6, E7 oncoproteins-induced events result in uncontrolled cell growth, accumulation of carcinogenic mutations, cell transformation, and tumor development [18].

p16INK4a overexpression is considered an excellent marker for infection with HPV-16 and 18 types. The tumor suppressor protein p16 functions as a Cyclin-dependent kinase inhibitor that inhibits Rb phosphorylation initiated by the cyclin D1–cyclin-dependent kinase complex. p16 can arrest the cell cycle in the G1 phase by regulating Rb phosphorylation [19-20]. It was reported that independent of head and neck cancer site, HNSCC cases positive for both p16 protein overexpression and HPV-16 DNA had 92 % increase in disease-specific survival [21].

Analysis of 229 metabolic genes between HPV-positive and negative HNSCC patients have revealed lower expression levels of genes associated with glycolysis and higher expression of genes involved in the tricarboxylic acid cycle, oxidative phosphorylation, and β-oxidation in HPV positive patient group compared to HPV-negative. SDHC, COX7A1, COX16, COX17, ELOVL6, GOT2, and SLC16A2 genes down-regulation improved patient survival in the HPV-positive patient group [22]. HPV positive oral squamous cell carcinoma (OSCC) is different from HPV negative OSCC on a histopathological and clinical level [23]. HPV-associated OSCC has a distinct pathogenesis and response to treatment [24]. The overall survival (OS) of HPV-positive OSCC is better than that of HPV-negative OSCC [12, 24].

The North-Eastern (NE) region of India is culturally as well as ethnically diverse with unique lifestyle and food habits which may have an important role in the complex interaction of environmental and genetic factors leading to a higher incidence of oral cancer [14]. There is insufficient evidence on the hr-HPV status and p16 protein expression with 5-year survival in OSCC patients of NE India. Our study aimed to investigate the 5-year survival of OSCC patients of NE India with hr-HPV status along with p16 protein expression.

## MATERIALS AND METHODS

### Patient Selection, Specimen Collection and Genomic DNA Extraction

A total of 72 histologically confirmed OSCC patients with written informed consent were enrolled in the study between January 2011 and January 2014 at Dr. Bhubaneswar Borooah Cancer Institute. Biopsy tissue specimens of the enrolled patients were collected before treatment. In all cases, multimodality treatment was given with a curative intent and the patients were followed up until December 2019. Participants were interviewed using a predesigned questionnaire to collect information about demographic and epidemiologic characteristics such as age, gender, tobacco usage etc whereas clinical characteristics including histology, stage, grade, treatment, and survival outcome were recorded during the course of follow-up. The study was approved by the institutional ethics committee of the Dr. Bhubaneswar Borooah Cancer Institute (BBCI) [BBCI/IEC-21/10]. Tissue biopsy specimens were collected in RNAlater (Qiagen, Germany) and stored at -80°C until further use.

Tissue specimens (25-50mg) were homogenized in 200µL volume of sterile phosphate-buffered saline (PBS) by using handheld plastic micro-pestle in ((PBS).PBS).Genomic DNA was extracted according to manufacturer’s instructions using QIAamp DNA mini kit (Qiagen, Germany). Bio-photometer Plus Nano-spectrophotometer (Eppendorf, Germany) was used for measuring genomic DNA quantity and quality at A260 nm and A280 nm. Quality was also determined using 1% Agarose gel electrophoresis (Amresco, USA). All specimen extracted genomic DNA has ratio of 1.7-1.8 at 260 nm / 280 nm [25].

### E6/E7 nested multiplex PCR (NMPCR) for HPV genotyping

For HPV detection and genotyping, HPV E6/E7 nested-PCR and HPV-type specific nested multiplex primer sequences were used as described previously [14, 26]. The Nested PCR protocol consisted of two rounds of PCR, and the PCR product of the E6/E7 served as the template for the multiplex hr-HPV typing (HPV-16, -18, -31, -45, 59). PCR reactions were carried out in a final volume of 20 µl of the PCR master mixtures using 25–100 ng genomic DNA, 1x Maxima HotStart PCR buffer with 1.5 mM MgCl_2_ (Thermo Scientific, USA), 250 nM of each primer (Metabion, Germany), 250 μM of each dNTPs (Thermo Scientific, USA) and 0.5 U Maxima Hot Start Taq DNA Polymerase (Thermo Scientific, USA).

Nested Multiplex PCRs (NMPCRs) were performed under the following cycling conditions for 1^st^ round of E6/E7 PCR (630bp): initial denaturation at 94 °C for 4 min, 40 cycles of 94 °C for 30 s, 52 °C for 45 s and 72 °C for 1 min 30 s, and final elongation at 72 °C for 7 min.

2^nd^ round of nested PCR reaction for HPV genotyping using multiplex HPV type-specific primers (Table 1) were done under the following cycling conditions: initial denaturation at 94 °C for 4 min, 40 cycles of 94 °C for 30 s, 56 °C for 30 s and 72 °C for 30 s, and final elongation at 72 °C for 7 min. B-globin as an internal positive control, HPV positive control containing confirmed HPV positive cervical cancer tissue DNA, and no template control using sterile deionized water were incorporated in each PCR reaction [14]. All samples of hr-HPV positive and HPV negative outcomes were reconfirmed by random sampling & testing by another investigator blinded about the outcome. The PCR products were analyzed by 2% agarose gel (Amresco, USA) containing ethidium bromide (Amresco, USA) visualized under in Gel Doc XR™ system (Bio-Rad, USA) (Fig. 2) [27].

**Table 1:**
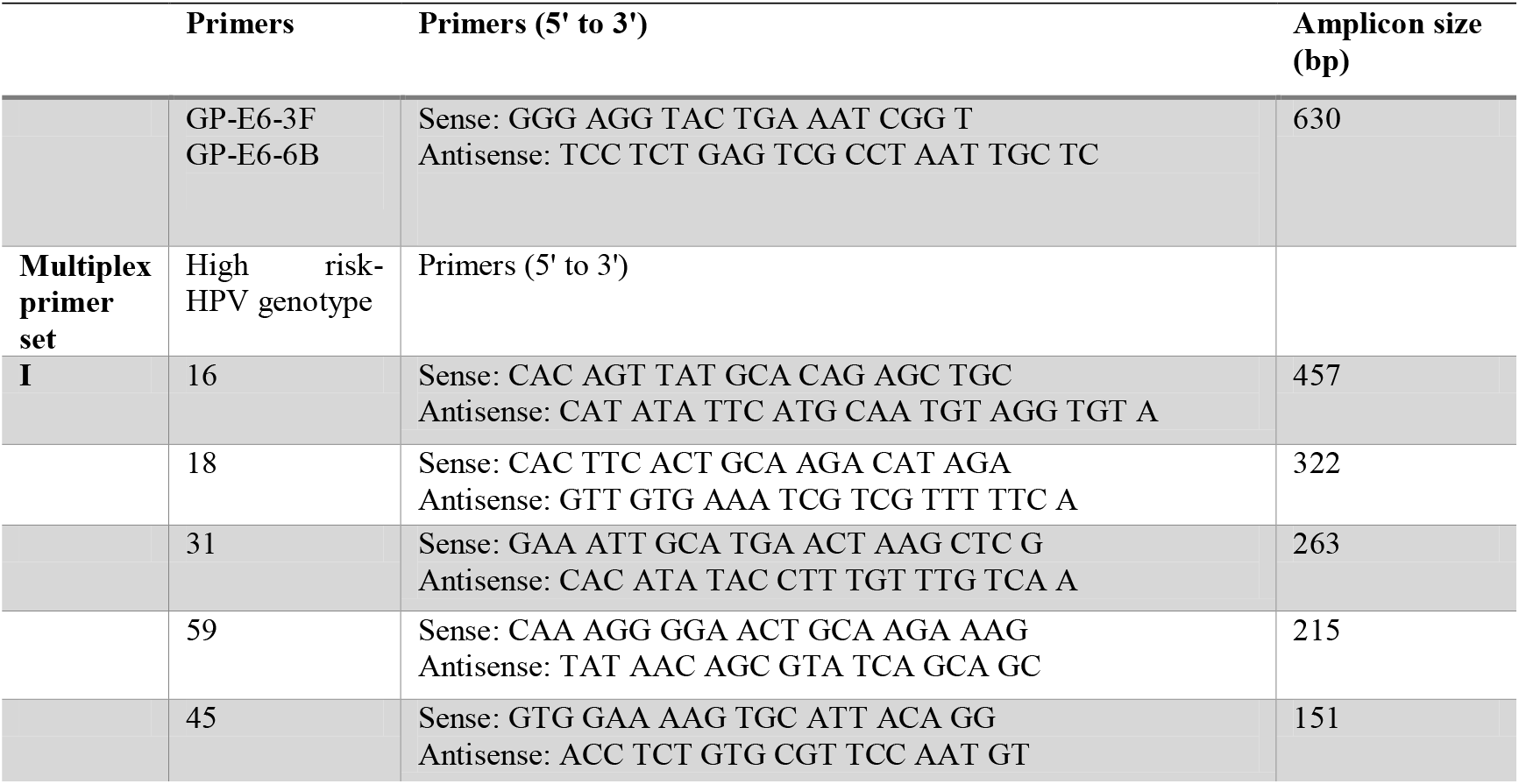
hr-HPV type primers specifications and amplicon size

**Figure 1:**
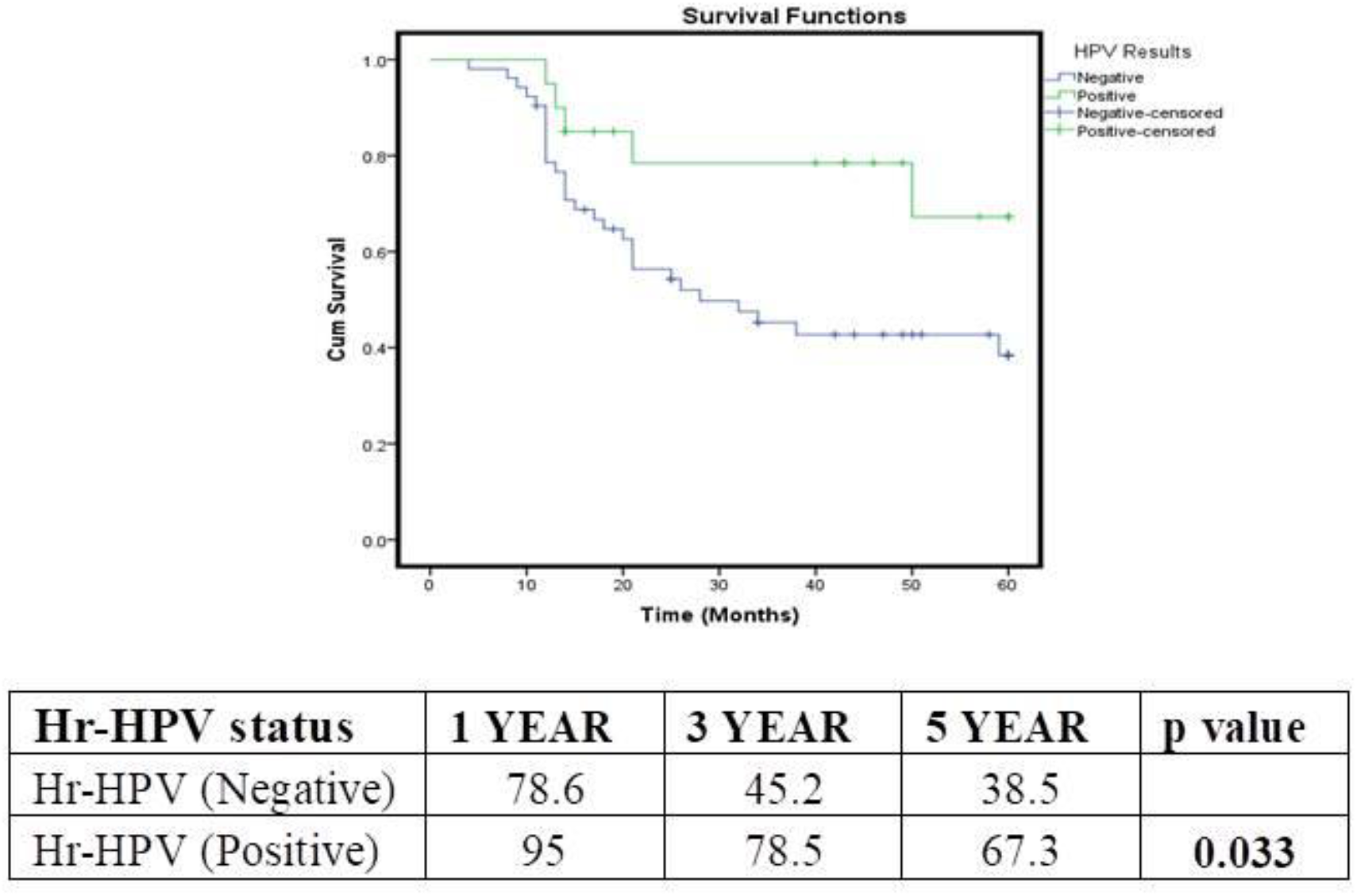
Kaplan–Meier Survival Curve of hr-HPV positive and negative OSCC Patients

**Figure 2:**
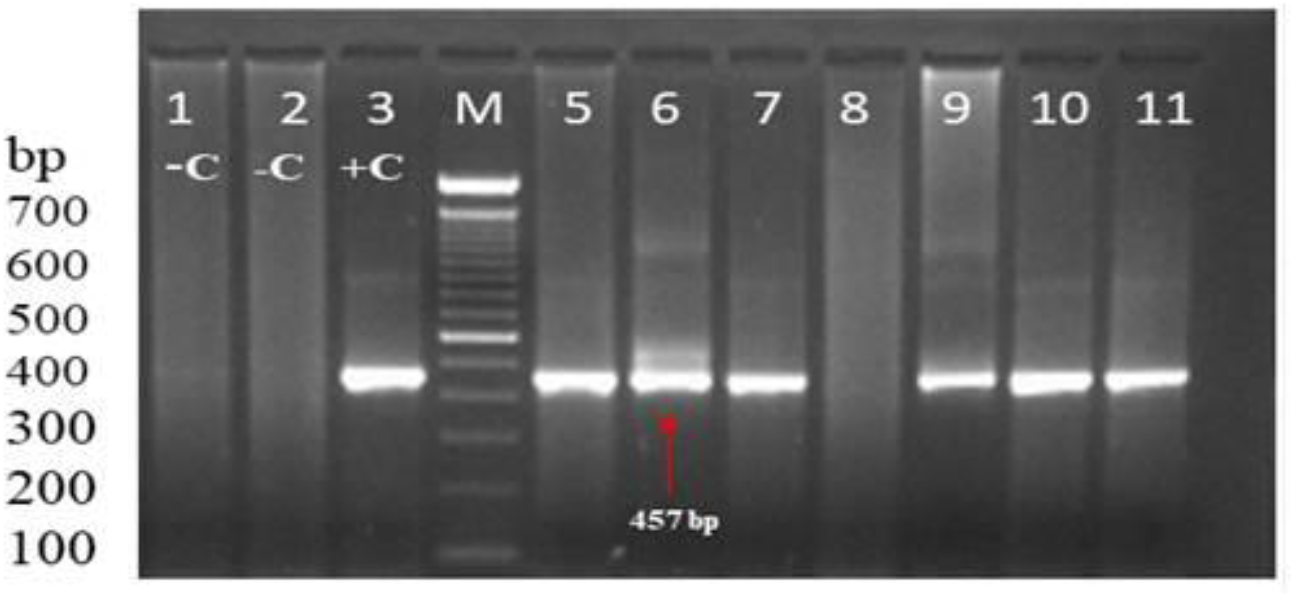
Representative image of agarose gel (2%) showing HPV Type-specific E6 Nested Multiplex PCR (NMPCR) Product. Lane 1, 2-negative control, Lane 3 – positive control for cervical cancer of M= 100 bp DNA Ladder, Lane 5, 6, 7 and 9-11 showing hr-HPV-16 types (457 bp) detection in oral cancer cases.

**Figure 3:**
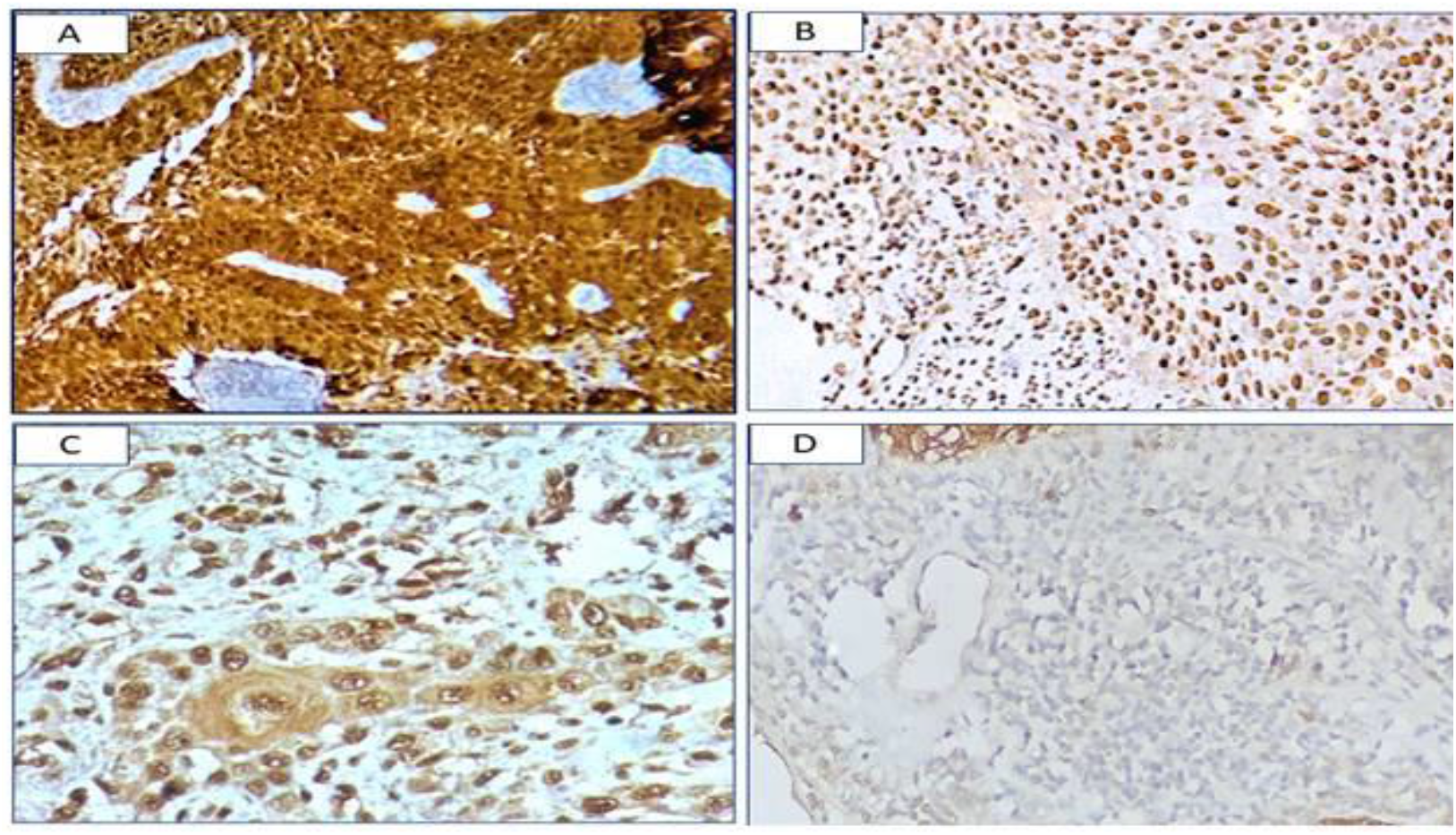
Immnunohistochemistry analysis for p16 protein expression. (A) high expression of p16 detected in both nuclei and cytoplasm in oral tumor. (B) moderate expression of p16 signal in both nuclei and cytoplasm (C) low expression with a clear positive nuclear but less cytoplasmic in tumour cells. (D) p16 low expression in both the nuclei and cytoplasm. Original 1nagnificatio11: A-D − ×400.

### Immunohistochemical p16 staining

As part of a standard procedure, the same malignant tumor specimen used for HPV genotyping was used for immunohistochemistry evaluation of pl6INK4a. All tissues were fixed in neutral buffered formalin before paraffin embedding, which is a standardized procedure. Briefly, formalin fixed paraffin-embedded (FFPE) sections were sliced (5 µm thickness) and fixed on poly L-lysine coated slides. The fixed sections were deparaffinized, rehydrated, and incubated in methanol containing 10 % H_2_O_2_ to reduce the endogenous peroxide activity. Antigen retrieval was performed in citrate buffer (pH 6.0) in the microwave and then the sample was kept in blocking buffer solution (1X PBS + 30X H202). The sample was incubated at 4°C overnight with mouse monoclonal p16INK4A antibody (BioGenex, USA) diluted (1:50) in 20 mM Tris-buffered saline (pH 7.4) containing 2mM CaCl_2_ solution and bovine serum albumin (BSA), and the sections were visualized using high sensitivity peroxidase DAB system (Dako REAL™ EnVision™ Detection System, Glostrup, Denmark), and counterstaining done with hematoxylin. All the tissue sections were then dehydrated by incubation in ascending ethanol series (50%, 70%, 90%, and 100%). We used Cervical cancer FFPE tissue specimen as a positive control for p16 expression, and in the negative control, the primary antibody was absent. Stained tumor sections were analyzed by light microscopy (Olympus IX83, Japan), and images were captured at 40X magnifications. Both nuclear and cytoplasmic staining was considered for immunoreactivity (fig. 2-4). Intensity of staining was scored as 0+ (no staining), 1+ (weak staining), 2+ (moderate staining), 3+ (strong staining) [28].

### Treatment Modality

All OSCC patients enrolled in this study had performance status score that permitted curative intent therapy. Treatment recommendations were decided at the tumor board attended by faculties of Head and Neck Surgery, Radiation Oncology, Medical Oncology, Radiology, and Pathology. Surgery was performed as per disease classification guideline. Patients received concurrent chemoradiotherapy (CRT). Definitive Radiotherapy (RT) was delivered at 70 Gy in 35 fractions of 2 Gy. Most of the patients received weekly cisplatin chemotherapy at 30 mg/m2. Patients were followed up to 84 months (max) and assessed for treatment outcome as per RECIST 1.1 criteria for solid tumors.

### Statistical Analysis

All OSCC patient data associated with clinicopathological characteristics, hr-HPV status, p16 protein expression were employed using the chi-square test or Fisher’s exact test. Univariate analysis was done by calculating odds ratios (ORs) along with 95% confidence intervals (CIs) to summarize data outcome. We also analyzed 5-year overall survival (OS) of OSCC patients. Year-wise survival status in hr-HPV positive and negative OSCC was calculated using the Kaplan–Meier method. Univariate analysis was used to select independent prognostic factors effect of all parameters of interest (HPV status, p16 protein expression, 5 years survival status, and clinical parameters). The statistical significance was considered for p-value ≤ 0.05. All univariate and survival data estimates were performed using SPSS statistical software (version20).

## RESULTS

Table 2 summarizes 72 OSCC patient characteristics categorized into two groups based on 5-year survival. Between OSCC patients with <50years and >50years age, 5-year survival has a marginal difference. Compared with female OSCC patients, male OSCC patients have a 2.32 times better probability of five-year survival (OR-2.32 CI 95%: P-0.11). TNM Stage III OSCC patients have a 7.80 times better likelihood of five-year survival than stage IV patients (OR-7.80 CI95%; P-0.03). Squamous cell carcinoma was the majority histological type compared to the adenocarcinoma (5.27%). The 5-year survival rate for patients with moderately differentiated squamous cell carcinomas (MDSCC) is 2.41 times higher than the rate for other morphological types but the difference is not statistically significant (OR-2.41 CI-95%; P-0.31).

**Table 2:** Characteristics of OSCC patients overall Survival (OS) at 5th Year after Diagnosis and treatment

|  | All N=72 (%) | Overall survival 5-year N=38 (%) | overall survival <5-year N= 34 (%) | OR [95%CI] Unadjusted OR | P- value |
| --- | --- | --- | --- | --- | --- |
| **Age** |  |  |  |  |  |
| <50 Years | 29 (40.27) | 15 (39.47) | 14 (41.17) | **1 (References)** |  |
| ≥50 Years | 43 (59.73) | 23 (60.53) | 20 (58.83) | 1.07 [ 0.41-2.75] | 0.88 |
| **Gender** |  |  |  |  |  |
| Female | 2 (29.16) | 8 (21.05) | 13 (38.23) | **1 (References)** |  |
| Male | 51 (72.83) | 30 (78.95) | 21(61.76) | 2.32 [0.81- 6.58] | 0.11 |
| **Stage** |  |  |  |  |  |
| II | 11(15.27) | 5 (13.15) | 6 (17.64) | **1 (References)** |  |
| III | 15 (20.83) | 13 (34.21) | 2 (5.88) | 7.80[1.16-52.35] | **0.03\*** |
| IV | 46 (63.89) | 20 (52.63) | 26 (76.47) | 0.93 [0.24-3.46] | 0.90 |
| **Histology** |  |  |  |  |  |
| Adenocarcinoma Carcinoma | 4 (5.66) | 2 (5.27) | 2 (5.99) | **1 (References)** |  |
| Squamous Cell Carcinoma | 68 (94.44) | 36 (94.73) | 32 (94.11) | 1.12 [0.14- 8.45] | 0.90 |
| **Morphology** |  |  |  |  |  |
| Well Differentiated † | 61(84.72) | 31 (81.57) | 30 (88.24) | **1 (References)** |  |
| Moderately Differentiated | 7 (9.59) | 5 (13.15) | 2 (5.88) | 2.41 [0.43- 13.44] | 0.31 |
| Poorly Differentiated | 4 (5.55) | 2 (5.26) | 2 (5.88) | 0.96 [0.12-7.31] | 0.97 |

**\*Statistically Significant**

### Hr-HPV status and survival analysis Outcomes

The 5-year unadjusted survival for hr-HPV positive & HPV negative OSCC and Kaplan – Meier (KM) estimates of survival curves for 1st year to 5th year are shown in (Table 3) and (Fig. 1), respectively. Among (n=20) hr-HPV positive OSCC patients, ≥ 5year survival was 80% (n=15) and <5 years survival was 20% (n=5) whereas among hr-HPV negative patients (n=52), ≥ 5year survival was 44.23% (n=23) and <5-year survival was 55.76% (n=29) respectively. We found the median survival was significantly higher for hr-HPV positive OSCC patients for 1st year (95%), 3 years (78.5%) and 5 years (67.3%)] as compared to median survival of hr-HPV negative OSCC patients for 1st year (78.6%), 3 years (45.2%) and 5 years (38.5%) [P-value = 0.03] (Fig1).

**Table 3:** Comparison of Overall 5 Year survival between hr-HPV Positive and HPV negative OSCC patients

|  | hr-HPV Positive |  |  |  |  | HPV Negative |  |  |  |  |
| --- | --- | --- | --- | --- | --- | --- | --- | --- | --- | --- |
|  | Total<br>N=20<br>(%) | Overall survival<br>5-year N= 15<br>(%) | Overall survival<br><5 years<br>N=5 (%) | OR [95%CI] | P- value | Total<br>N=52 | Overall<br>survival 5-year N=23<br>(%) | Overall survival<br><5 years<br>N=29 (%) | OR [95%CI] | P- value |
| **Age** |  |  |  |  |  |  |  |  |  |  |
| <50 Years | 9(45) | 6 (40) | 3 (60) | **1 (References)** |  | 19 (36.54) | 8 (34.79) | 11 (37.93) | **1 (References)** |  |
| ≥ 50 Years | 11 (55) | 9 (60) | 2 (40) | 2.25 [0.28-17.76] | 0.44 | 33 (63.46) | 15 (65.21) | 18 (62.06) | 1.14 [0.36- 3.58] | 0.81 |
| **Gender** |  |  |  |  |  |  |  |  |  |  |
| Female | 5 (25) | 2 (13.33) | 3 (60) | **1 (References)** |  | 16 (30.10) | 6 (26.08) | 10 (34.48) | **1 (References)** |  |
| Male | 15 (75) | 13 (86.66) | 2 (40) | 9.75 [0.95- 99.96] | **0.05\*** | 36 (69.23) | 17 (73.91) | 19 (65.51) | 1.9 [0.44-4.97] | 0.51 |
| **TNM Staging** |  |  |  |  |  |  |  |  |  |  |
| II | 2 (10) | 1 (6.66) | 1(20) | **1 (References)** |  | 9 (17.31) | 4 (17.39) | 5 (17.24) | **1 (References)** |  |
| III | 2 (10) | 2 (13.33) | 0 (0.0) | - 0.4 |  | 13 (25) | 11(47.82) | 2 (6.89) | 6.87 [0.93- 50.78] | **0.05\*** |
| IV | 16 (80) | 12 (80) | 4 (80) | 3.00 [0.15-59.89] | 0.47 | 20 (38.46) | 8 (34.79) | 22 (75.86) | 0.45 [0.09-2.12] | 0.31 |
| **Histology** |  |  |  |  |  |  |  |  |  |  |
| Adenocarcinoma<br>Carcinoma | 0 | 00 | 0 (0.0) | **1 (References)** |  | 4 (7.70) | 2 (8.69) | 2 (6.89) | **1 (References)** |  |
| Squamous Cell<br>Carcinoma | 20 (100) | 15 (86.67) | 5 (100) | - | 0.61 | 48 (92.30) | 21 (86.96) | 27 (86.20) | 0.77 [0.10-5.98] | 0.80 |
| **Morphology** |  |  |  |  |  |  |  |  |  |  |
| **Well**<br>**Differentiated** | 16 (80) | 12 (80) | 4 (80) | **1 (References)** |  | 45 (86.58) | 19 (82.60) | 26 (89.65) | **1 (References)** |  |
| Moderately<br>Differentiated | 3 (15) | 2 (13.33) | 1 (20) | 0.76 [0.04- 9.47] | 0.76 | 4 (7.70) | 3 (13.04) | 1 (3.44) | 4.10 [0.39- 42.5] | 0.23 |
| Poorly<br>Differentiated | 1 (5) | 1 (6.66) | 0 (0.0) | - 0.96 |  | 3 (5.80) | 1 (4.34) | 2 (6.89) | 0.68[0.05- 8.10] | 0.76- |

**\*Statistically Significant**

Hr-HPV positive OSCC patients in ≥50 years age has 2.25 times better 5-year survival than the HPV negative group (OR-2.25 CI-95%; P-0.44). Male OSCC patients of the hr-HPV positive group have 9.75 times significantly better 5-year survival than the HPV negative group (OR-9.75 CI-95%; P-0.05). hr-HPV positive OSCC stage IV patients have 3.0 times better 5-year survival than the HPV negative group (OR-3.00 CI-95%; P-0.47). With respect to morphological types, there was no significant difference observed between hr-HPV positive and HPV negative group of patients.

### Association of P16 expression, hr-HPV status with 5 -year survival

Association of survival status of OSCC patients with P16 expression and hr-HPV status was described in (Table 4). We found that among ≥5-year survival OSCC patients, P16 protein overexpression was 28% (n=11), moderate expression 39.47% (n=15) and low expression 13.15 % (n=5) whereas among <5-year survival OSCC patient group, P16 protein overexpression was 26.47% (n=9), moderate expression 35.29% (n=12) and low expression 11.76 % (n=4) respectively. P16 expression (low to overexpression) group and p16 expression negative group of OSCC patients have not shown any significant association with 5-year survival.

**Table 4:**
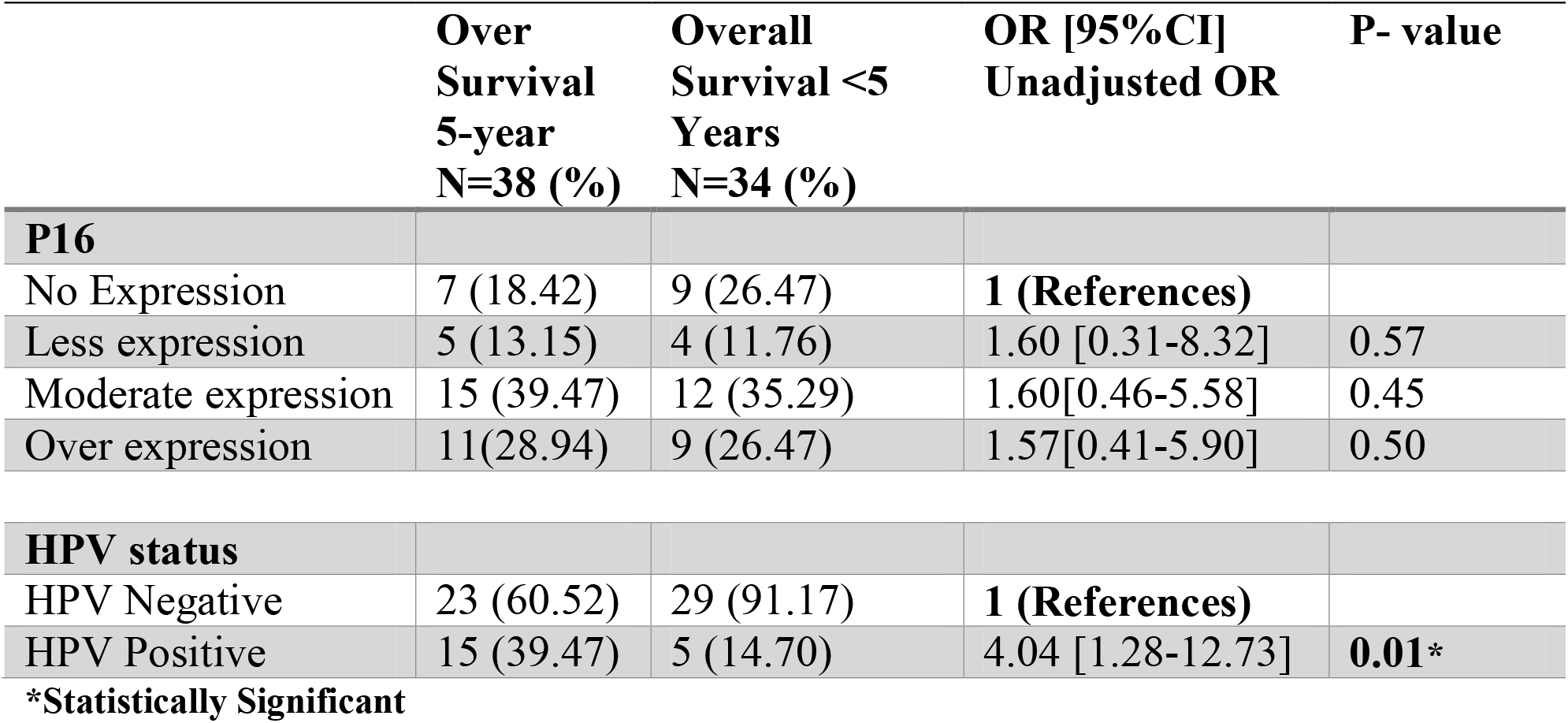
Overall Survival status of OSCC patients with reference to P16 expression and HPV status

|  | **Over**<br>**Survival**<br>**5-year**<br>**N=38 (%)** | **Overall**<br>**Survival <5**<br>**Years**<br>**N=34 (%)** | **OR [95%CI]**<br>**Unadjusted OR** | **P- value** |
| --- | --- | --- | --- | --- |
| **P16** |  |  |  |  |
| No Expression | 7 (18.42) | 9 (26.47) | **1 (References)** |  |
| Less expression | 5 (13.15) | 4 (11.76) | 1.60 [0.31-8.32] | 0.57 |
| Moderate expression | 15 (39.47) | 12 (35.29) | 1.60[0.46-5.58] | 0.45 |
| Over expression | 11(28.94) | 9 (26.47) | 1.57[0.41-5.90] | 0.50 |
| **HPV status** |  |  |  |  |
| HPV Negative | 23 (60.52) | 29 (91.17) | **1 (References)** |  |
| HPV Positive | 15 (39.47) | 5 (14.70) | 4.04 [1.28-12.73] | **0.01\*** |

**\*Statistically Significant**

hr-HPV group of OSCC patients has shown 4.04 times significantly better 5-year survival as compared to HPV negative group of patients (OR-4.04 CI95%; P-0.01).

## DISCUSSION

We found hr-HPV DNA positivity of 27.7% (n=20) in OSCC patients of North –East India. Tian et al. (2019), in 24000 HNSCC patients of the American population, found HPV-positivity by disease site-wise at 17.7% for Hypopharynx, 11% for larynx, 10.6% for Oral Cavity, and 62.9% for oropharynx [29]. They also found that HPV positivity was associated with superior overall survival (OS) in patients with AJCC stage III to IVB OSCC (HR, 0.78; P - 0.03). Widła et al. (2020) found 16.0% HPV DNA positivity in polish oral cancer patients. They also suggested that HPV DNA is an independent favorable prognostic factor for both OS and Disease-free survival (DFS) in oropharyngeal as well as in OSCC patients [18]. hr-HPV DNA positivity was found to be 17.9%, 30.8%, 7.0%, 3.3% in OSCC patients of Indoanesia, Bosnia, Canada, Switzerland respectively [30-33]. HPV DNA in OSCC patients was found to be in the range of 15-48% in studies from various regions of India [3,7, 34].

We found that in OSCC, hr-HPV prevalence and 5-year survival were higher in men. A similar outcome was observed in the US population-based National Health and Nutrition Examination Survey (NHANES) [35]. It was also reported that oral HPV infection to be more common in Caucasian women <30 years of age [36].

We found in our study that TNM stage III has significantly superior 5-year survival in hr-HPV positive compared to HPV negative OSCC patients. It was earlier reported that I and II stage OSCCs exhibited significantly better 5-year survival (73.3%) than stage III and IV OSCCs (41.8%). Multivariate analysis also suggested the tumor stage as a significant prognostic indicator of survival [37, 38]. Lin et al. (2020) found distinct survival differences between the early stage and advanced stages of tumors in Chinese OSCC patients were associated with recurrence [39]. Xu et al. (2018) observed that different histological grades of OSCCs had similar survival outcomes [40]. Early-stage was an independent prognostic factor for better survival in OSCC cancer patients but not for patients with advanced-stage disease [41].

We found that Male OSCC patients of the hr-HPV positive group have significantly better 5-year survival than the HPV negative group. A previous meta-analysis has shown that HPV-positive men have a better survival rate compared to HPV-positive women [42]. Ritchie *et al*. (2003) proposed that OSCC clinical path may have gender-associated survival differences. They found that men with HPV-positive tumors had a better prognosis than men with HPV-negative tumors, although no such association was observed in women [43]. However, some studies found that women have a significant survival advantage in several cancers other than head and neck cancer [44-46]. It was hypothesized that sex hormones might have a role in HPV-related cancers as a cofactor [47]. Progesterone antagonists and nuclease-resistant oligomers contain the HPV-16 response factor that has been shown to inhibit cell growth and transcription of the E6/E7 genes [47,48]. Survival advantage among women in OSCC may be partly explained by lower exposure to lifestyle factors such as alcohol and tobacco in Europe, Southeast Asia, and NE -India [15, 49-50].

Our study found that p16 expression has shown a trend of improved 5-year survival, but this association was not statistically significant. A Danish study has also shown no difference in survival outcome [51]. Early-stage OSCCs had higher levels of p16 expression, which was linked to more remarkable survival after surgery and radiotherapy [52]. hr-HPV infection was reported to be associated with an improved 5-year overall survival in German OSCC patients compared to HPV-negative OSCC patients [15]. It was also reported that HPV positivity had no association with overall survival for all sites of the oropharynx, hypopharynx, oral cavity, and larynx subsites [53].

HPV-AHEAD study conducted in Indian head and neck cancer patients found, hr-HPV DNA in 13.7% of HNC patient samples and 1.6% in oral cavity cancers. More than 50% of HPV DNA/RNA-positive cases were p16INK4a-negative, whereas 17.9% of HPV RNA-negative cases were p16INK4a-positive. They concluded that p16INK4a staining might not be an excellent surrogate marker of HPV in Indian HNC patients [7]. Our study limitations may be small sample size.

## CONCLUSION

Our study found that hr-HPV DNA positive status may be a significantly good indicator of better 5-year survival in OSCC patients of North-East India. p16 expression has not shown any significant association with improvement of 5-year survival. Further studies in large cohort of OSCC patients of North-East India may be required to validate the outcome of the present study for clinical practice.

## Data Availability

All authors declare that data of the research study are provided in tables and figures. There is no dataset/ software created as part of this study.

## ACKNOWLEDGEMENTs

The authors are grateful to the Department of Biotechnology (DBT), Government of India, New Delhi, India, research grant (No. BT/Med/NE-SFC/2009) for this study. The authors express their gratitude to all participants of this study.

## DECLARATIONS

### Code availability (software application or custom code)

**Not Applicable**

### Ethics Approval

Study was conducted in compliance of the Declaration of Helsinki as a statement of ethical principles for medical research involving human subjects, including research on identifiable human material and data. The study was approved by the institutional ethics committee of Dr. Bhubaneswar Borooah Cancer Institute (BBCI) [BBCI/IEC-21/10]. The Institutional ethics committee of Dr. Bhubaneswar Borooah Cancer Institute is registered (ECR/1040/Inst/AS/2018/RR-22) with Central Drugs Standard Control Organization, Directorate General of Health Services, Ministry of Health & Family Welfare, Government of India.

### Consent to Participate

All the OSCC patients of the study were enrolled after informed written consent.

### Consent for Publication

All authors have read and approved the manuscript for submission and publication.

